# In-host Modelling of COVID-19 in Humans

**DOI:** 10.1101/2020.03.26.20044487

**Authors:** Esteban A. Hernandez-Vargas, Jorge X. Velasco-Hernandez

## Abstract

COVID-19 pandemic has underlined the impact of emergent pathogens as a major threat for human health. The development of quantitative approaches to advance comprehension of the current outbreak is urgently needed to tackle this severe disease. In this work, several mathematical models are proposed to represent SARS-CoV-2 dynamics in infected patients. Considering different starting times of infection, parameters sets that represent infectivity of SARS-CoV-2 are computed and compared with other viral infections that can also cause pandemics.

Based on the target cell model, SARS-CoV-2 infecting time between susceptible cells (mean of 30 days approximately) is much slower than those reported for Ebola (about 3 times slower) and influenza (60 times slower). The within-host reproductive number for SARS-CoV-2 is consistent to the values of influenza infection (1.7-5.35). The best model to fit the data was including immune responses, which suggest a slow cell response peaking between 5 to 10 days post onset of symptoms. The model with eclipse phase, time in a latent phase before becoming productively infected cells, was not supported. Interestingly, both, the target cell model and the model with immune responses, predict that virus may replicate very slowly in the first days after infection, and it could be below detection levels during the first 4 days post infection. A quantitative comprehension of SARS-CoV-2 dynamics and the estimation of standard parameters of viral infections is the key contribution of this pioneering work.

## INTRODUCTION

Epidemics by infectious pathogens are a major threat to humankind. The year 2020 has uncovered one of the biggest pandemics in history, the novel coronavirus SARS-CoV-2 that was first reported in Wuhan, Hubei Province, China in December 2019. SARS-CoV-2 causes a severe, potentially fatal acute respiratory syndrome (COVID-19). Thus far, about 267013 confirmed cases and about 11201 deaths were reported worldwide [1]. While China has made a large effort to shrink the outbreak, COVID-19 has developed into a pandemic in 185 countries. Case numbers are alarming as the virus spreads in Europe, Iran, South Korea, and Japan. In fact, the pandemic epicentre changed to Europe on 13 of March 2020.

Coronaviruses can be found in different species of animals (*e.g*. bats and camels) and can evolve and infect humans by droplets from coughing or sneezing. Previous outbreaks to COVID-2019 were the Severe Acute Respiratory Syndrome (SARS-CoV), reported in Asia in February 2003 resulting in 8422 cases with a case-fatality rate of 11% [1]. Later, in 2012, the Middle East respiratory syndrome (MERS-CoV) was identified in Saudi Arabia and infected 2506 people, killing 862 between 2012 and 2020 [1]. Metagenomics studies previous to the COVID-19 outbreak envisaged the possibility of future threats due to the identification of several sequences closely related SARS-like viruses circulating in the Chinese bat populations [2, 3].

Unfortunately, no vaccine or antiviral drug is likely to be available soon. In fact, either monoclonal antibody or vaccine approaches have failed to neutralize and protect from coronavirus infections [3]. Therefore, individual behaviour (*e.g* early self-isolation and social distancing) as well as preventive measures such as hand washing, covering when coughing are critical to control the spread of COVID-19 [4]. Additionally to these measures, several travel restrictions and quarantines have taken place in many countries around the globe.

Epidemiological mathematical models have been developed to help policy makers to take the right decisions [4]. These have highlighted that social distancing interventions to mitigate the epidemic is a key aspect. There are many epidemiological unknowns with 2019-nCoV [4]. The case fatality rate for COVID-19 is about 0 3–1% [1]. However, adjusted estimation by [5] indicates that COVID-19 mortality rate could be as high as 20% in Wuhan. In its early stages, the epidemic have doubled in size every 7.4 days [6]. Moreover, the basic reproductive number was estimated to be 2.2 (95% CI, 1.4 to 3.9) [6]. Based on the relative long incubation period for COVID-19, about 5–6 days [1], Anderson *et al*. [4] suggested that might be considerable pre-symptomatic infectiousness.

While there are many mathematical models developed at epidemiological level for COVID-19, there is none so far at within-host level to understand SARS-CoV-2 replication cycle and its interactions with the immune system (Fig.1). Among several approaches, the target cell model has served to represent several diseases such as HIV [7–10], Hepatitis virus [11, 12], Ebola [13, 14], influenza [15–18], among many others. A detailed reference for viral modelling can be found in [19]. Very recent data from infected patients with COVID-19 has enlighten the within-host viral dynamics. Zou *et al*. [20] presented the viral load in nasal and throat swabs of 17 symptomatic patients. Interestingly, SARS-CoV-2 replication cycles may last longer than flu, about 10 days or more after the incubation period [4, 20]. Here, we contribute to the mathematical study of SARS-CoV-2 dynamics at within-host level based on data presented by Wolfel *et al*. [21].

**Fig 1.**
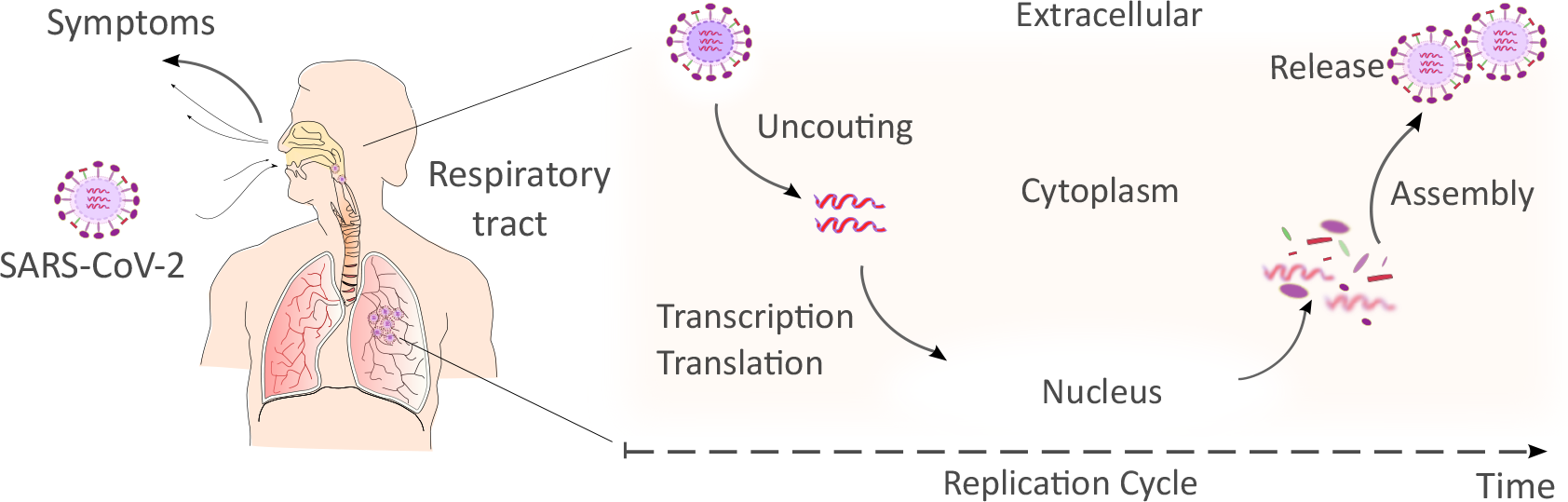
SARS-CoV-2 Replication Cycle. After the binding to receptors of the host cell, the virus RNA is uncoated in the cytoplasm. Then, transcription/translation processes take place to generate new viral RNA material and proteins. Virus assembly occurs within vesicles followed by virus release. Once the virus is released can infect other cells.

## RESULTS

Using ordinary differential equations (ODEs), different mathematical models are presented to adjust the viral kinetics reported by Woelfel *et al*. [21] in patients with COVID-19. Viral load [21] was sampled from throat swab cultures and measured in *Copies/mL, g Swab*, at Log10 scale. To understand the SARS-CoV-2 dynamics observed in infected patients, mathematical models are employed as both a quantitative recapitulation of experimental data and as a tool to prioritize mechanisms on the basis of mathematical models and the Corrected Akaike Information Criterion (AICc) for model selection. The cost function (14) is minimized to adjust the model parameters based on the Differential Evolution (DE) algorithm [22].

### Exponential Growth and Logarithmic Decay Model

Based on the experimental data [21], the viral dynamic is divided into two parts, exponential growth (*V*_*g*_) and decay (*V*_*d*_) modelled by equations (1) and (2), respectively.

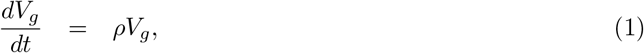

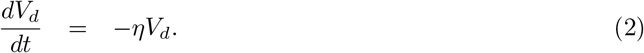

Viral growth is assumed to start at the onset of symptoms, with initial viral concentration *V*_*g*_(0). The parameter *ρ* is the growth rate of the virus. The parameter *η* quantifies the decay rate of the virus, while *V*_*d*_(0) the initial value of the virus in decay phase. Note that the growth phase of the virus was measured only in two patients (A, and B) [21].

Simulation are shown in Fig.2 and numerical results are presented in Table 1. The mean growth rate (*ρ*) is estimated as 3.98 (1/day) while the initial condition estimate is approximately 0.31 (Copies/mL). The mean decay rate of the virus (*η*) is around 0.95 (1/day), with the slowest rate estimate of 0.63 (1/day) presented for patients B, E, and F. The fastest decay rate was presented in the patient I with an estimate of 2.51 (1/day). This slow decay rate may explain the long duration of the virus (11-22 days) observed in the patients after the onset of symptoms [21].

**Fig 2.**
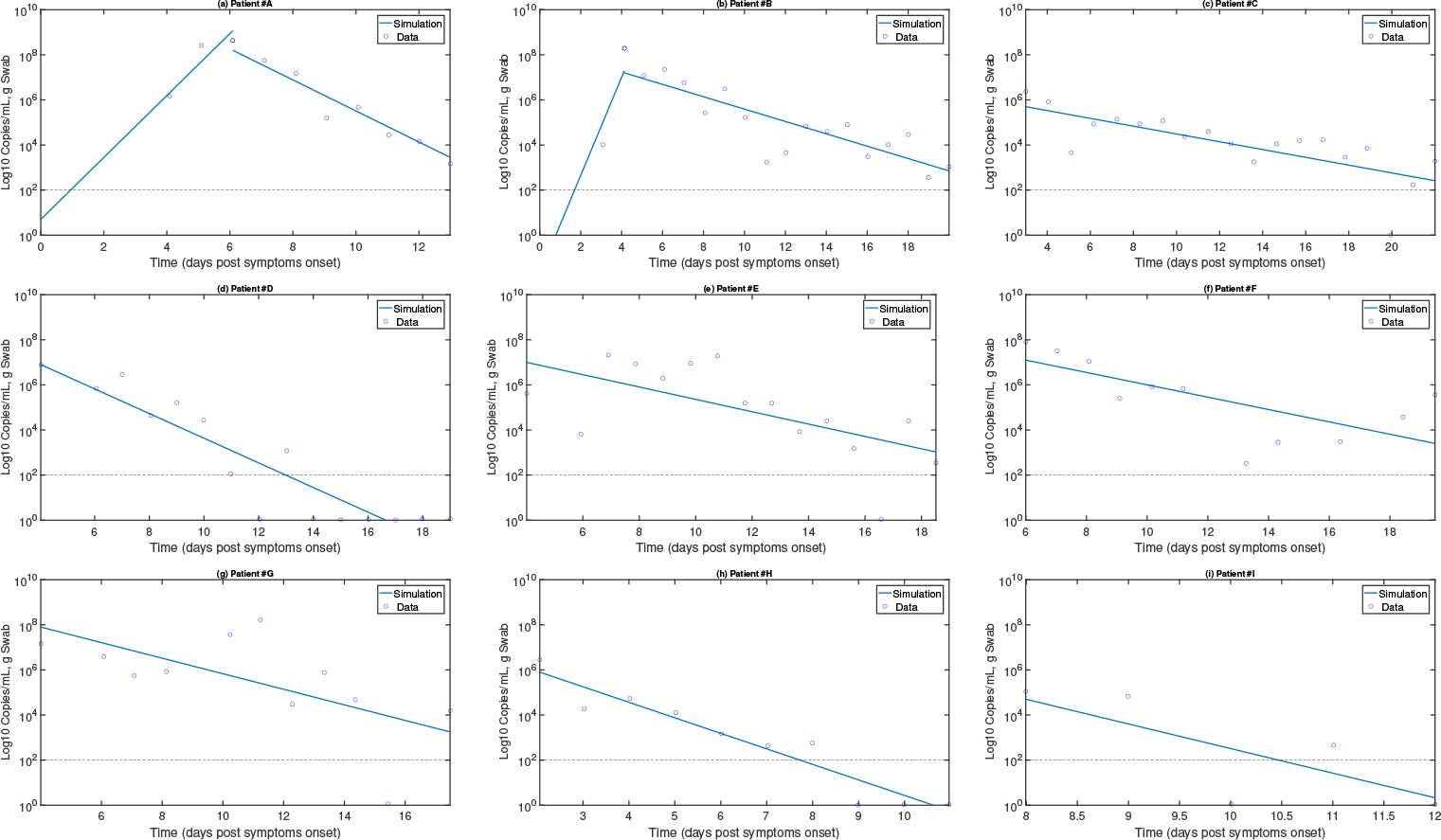
Exponential growth and decay model for COVID-19. Continuous line are simulation based on (1) for viral exponential growth (*V*_*g*_) or on (2) for viral decay (*V*_*d*_). blue circles represents the data from [21]. Viral growth rate (*ρ*) was only computed for patients A (till day 6) and B (till day 4) while the rest of patients have missing these measurements. For all patients viral decay rate *η* in (2) is computed.

**Table 1.** Estimations for the model (1)-(2) using experimental data from [21]. For the exponential growth phase there were measurements only for patient A and B, for the rest of patients were more in the logarithmic decay phase. This is the reason why patient A and B are the only ones that report estimations of viral growth.

| Patient | Growth |  | Decay |  |
| --- | --- | --- | --- | --- |
| | $\rho$<br>(1/day) | $V_g(0)$<br>(Copies/mL) | $\eta$<br>(1/day) | $V_d(0)$<br>(Copies/mL) |
| A | 3.16 | 5.01 | 1.58 | 8.20 |
| B | 5.01 | 0.02 | 0.63 | 7.20 |
| C |  |  | 0.39 | 5.7 |
| D |  |  | 1.26 | 6.9 |
| E |  |  | 0.63 | 7 |
| F |  |  | 0.631 | 7.1 |
| G |  |  | 0.79 | 7.9 |
| H |  |  | 1.58 | 5.9 |
| I |  |  | 2.51 | 4.7 |
| Mean | 3.98 | 0.31 | 0.95 | 6.64 |
| [Min-Max] | [3.16-5.01] | [0.02-5.01] | [0.39-2.51] | [4.7-8.21] |

### Target Cell Model

The mathematical model used here to represent coronavirus dynamics is based on the target cell-limited model [19, 23, 24]. Coronavirus can replicate in a variety of cell types, including epithelial cells. The coronavirus infection model is as follows:

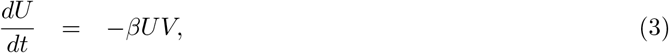

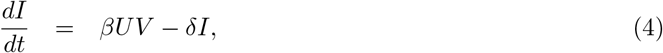

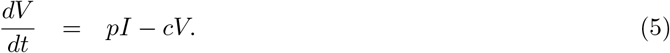

Host cells can be in one of following states: susceptible (*U*) and infected (*I*). Viral particles (*V*) infect susceptible cells with a rate *β* ((Copies/mL)^− 1^ day^− 1^). Once cells are productively infected, they release virus at a rate *p* (Copies/mL day^− 1^ cell^− 1^) and virus particles are cleared with rate *c* (day^− 1^). Infected cells are cleared at rate *δ* (day^− 1^) as consequence of cytopathic viral effects and immune responses.

Coronaviruses infect mainly in differentiated respiratory epithelial cells [25]. Previous mathematical model for influenza [17] have considered about 10^7^ initial target cells (*U* (0)). Initial values for infected cells (*I*(0)) are taken as zero. *V* (0) is determined from estimations in Table 1. Note that *V* (0) cannot be measured as it is below detectable levels (about 100 *Copies/m*) [21].

Viral kinetics are measured after the on-set of symptoms [21], however, it is unknown when the initial infection took place. Patients infected with MERS-CoV in [26] showed that the virus peaked during the second week of illness, which indicated that the median incubation period was 7 days (range, 2 to 14) [26]. For parameter fitting purposes, we explore three different scenarios of initial infection day (*t*_*i*_), that is, -14, -7, -3 days before the onset of symptoms for patients A and B, see Fig. 3.

**Fig 3.**
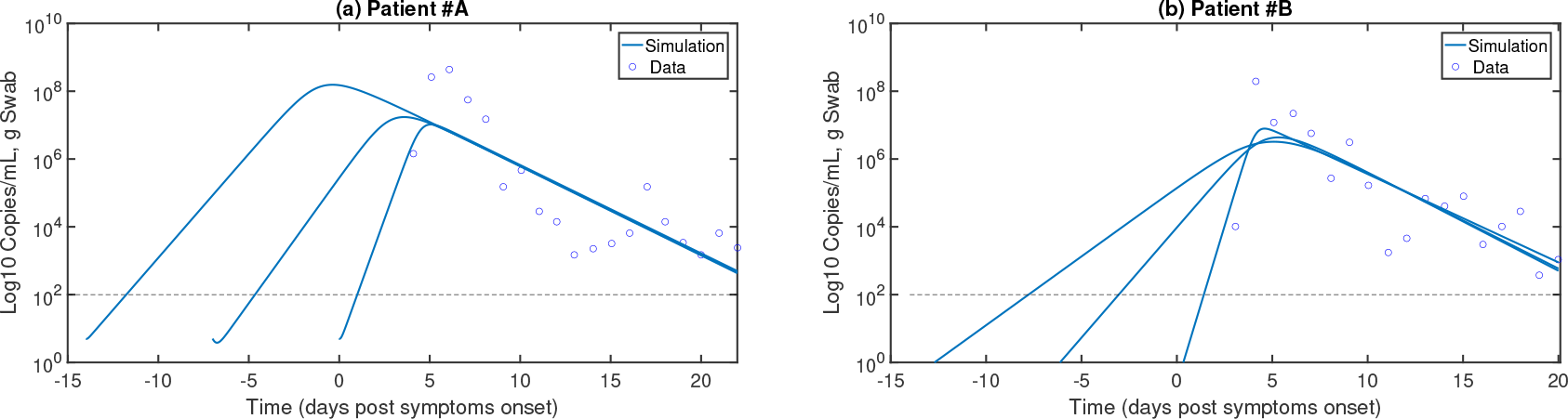
Target cell model for SARS-CoV-2. Continuous line are simulation based on the taget cell model (3)-(5). Blue circles represents the data from [21]. Due to the most complete data sets in [21] were from patient A and B, then these are the only presented in panel (a) and (b), respectively. Infection time was assumed at -14, -7 and 0 days post symptom onset.

Infectivity can be defined as the ability of a pathogen to establish an infection [27]. To quantify infectivity, the within-host reproductive number (*R*_0_) was computed. *R*_0_ is defined as the expected number of secondary infections produced by an infected cell [28]. When *R*_0_ < 1, one infected individual can infect less than one individual. Thus, the infection would be cleared from the population.

Otherwise, if *R*_0_ > 1, the pathogen is able to invade the target cell population. This epidemiological concept has been applied to the target cell model (3)-(5), with

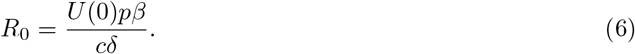

Previous studies [13, 29, 30] provided estimates of the *infecting time* (*t*_inf_), that represents the time required for a single infectious cell to infect one more cell. Viruses with a shorter infecting time have a higher infectivity [29, 30]. From equations (3)-(5), *t*_inf_ can be explicitly computed as:

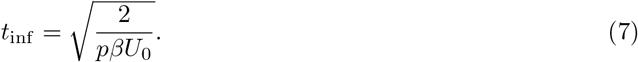

Assuming day of infection at day 0 post symptom onset (pso) would result in very high reproductive numbers (*R*_0_) and a high infection rate (*β*) for patients A and B as presented in Table 2. Alternatively, assuming the initial day of infection is either day -14 or -7 pso, then the rate of infection of susceptible cells (*β*) would be slow but associated with a high replication rate (*p*).

**Table 2.** Estimations for the target cell model (3)-(5) using experimental data from [21] for patients A and B.

| Patient | $t_i$<br>(days) | $\beta$ | $\delta$ | $p$ | $c$ | $R_0$ | $t_{\text{inf}}$<br>(hours) | AIC |
| --- | --- | --- | --- | --- | --- | --- | --- | --- |
| A | 0 | $3.97 \times 10^{-7}$ | 4.71 | 8.2 | 0.6 | 11.5 | 32.6 | 12.00 |
| | -7 | $9.98 \times 10^{-8}$ | 0.61 | 9.3 | 2.3 | 6.6 | 61.2 | 12.90 |
| | -14 | $5.00 \times 10^{-9}$ | 11.01 | 525 | 0.7 | 3.4 | 36.4 | 12.99 |
| B | 0 | $5.61 \times 10^{-7}$ | 11.1 | 13.4 | 0.6 | 11.3 | 21.5 | 5.99 |
| | -7 | $1.77 \times 10^{-7}$ | 14.11 | 20.2 | 0.8 | 3.17 | 31.12 | 10.78 |
| | -14 | $7.06 \times 10^{-8}$ | 58.31 | 195.8 | 1.4 | 1.7 | 15.85 | 12.15 |

Strikingly, Fig. 3(b) reveals a long period (about 4 days post infection) of viral replication below detectable levels. Independently of the starting infection time (*t*_*i*_), numerical results at the Table 2 reveal very consistent reproductive numbers for patients A and B (approximately 11),implying that SARS-CoV-2 would invade most of the susceptible target cells. Remarkably, the infecting time *t*_inf_ is slow, about 30 hours. This may explain why SARS-CoV-2 can last several days (12-22 days pso) in infected patients [21].

### Target Cell Model with Eclipse Phase

To represent the time frame of the infection more adequately, an additional state is added where newly infected cells spend time in a latent phase (*E*) before becoming productively infected cells (*I*) [29, 31]. This can be written as follows:

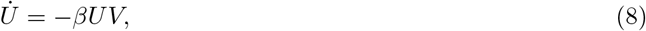

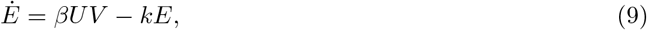

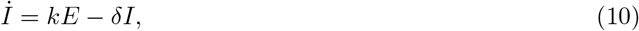

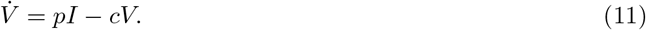

Cells in the eclipse phase (*E*) can become productively infected at rate *k*. Holder *et al*. [29] considered different time distributions for the eclipse phase and viral release by infected cells for influenza. Their results showed that the time distribution of the eclipse phase and viral release directly affect the parameter estimation. For SARS-CoV-2, Fig.4 the eclipse phase model (AIC≈34) does not improve the fitting respect to the target cell model (Table 2) even when very long eclipse phase periods are assumed (*e.g* 100 days), implying that this mechanism could be negligible on SARS-CoV-2 infection.

**Fig 4.**
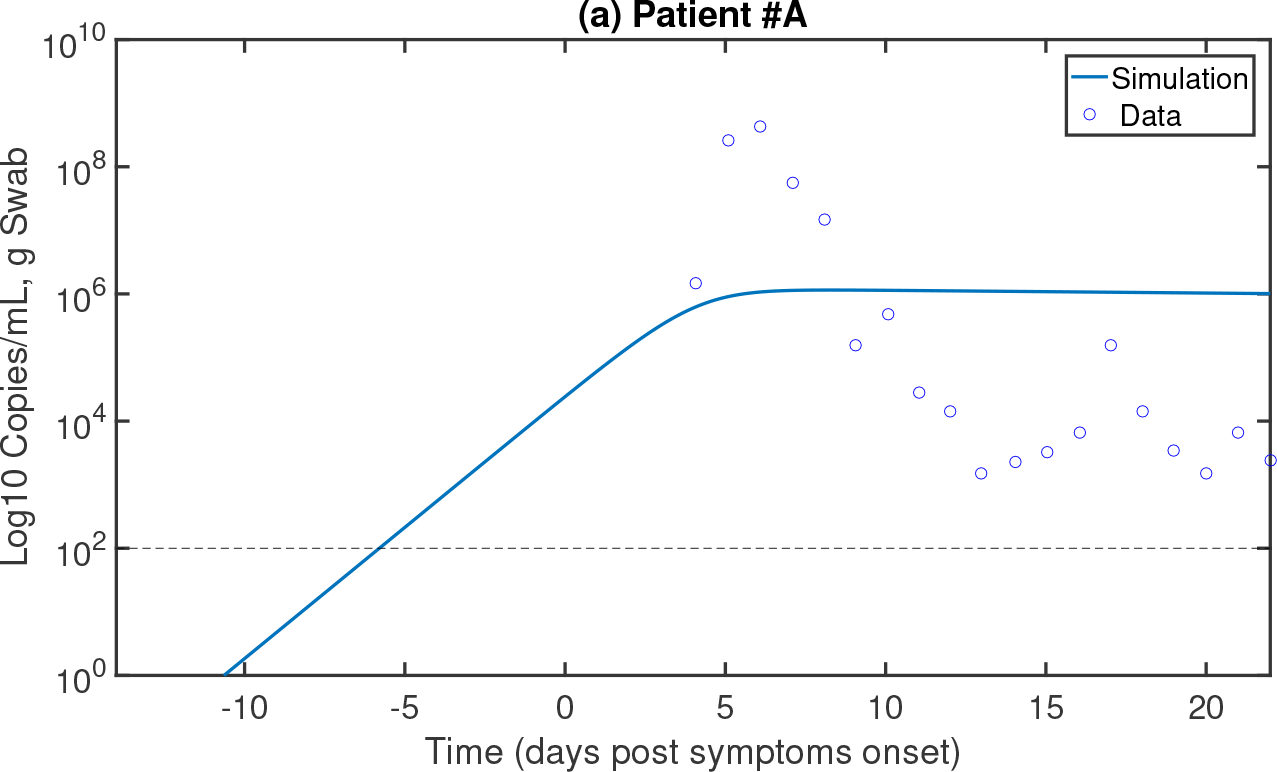
Target cell model with eclipse Phase for SARS-CoV-2. Continuous line are simulation based on the target cell model (8)-(11). The parameter *k* was fix to 0.01 day^− 1^. Blue circles represents the data from [21]. The hypothesis of eclipse phase during COVID-19 is not supported as it has a higher AIC value (approximately 34) than the target cell model. Infection time was assumed at -14 days post symptom onset.

### Mathematical Model with Immune Response

Previous studies have acknowledged the relevance of the immune T-cell response to clear influenza [17, 32–36]. Due to identifiability limitations for the estimation of the parameters of the target cell model using viral load data, a minimalistic model was derived in [37, 38] to represent the interaction between the viral and immune response dynamics. The model assumes that the virus (*V*) level induces the proliferation of T cells (*T*) as follows:

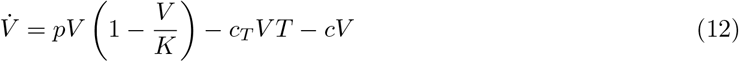

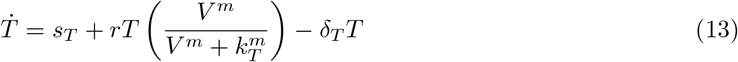

Viral replication is modelled with a logistic function with maximum carrying capacity *K* and replication rate *p*. The virus is cleared at a rate *c*. The term *c*_*T*_ *V T* represents the rate of killing of infected cells by the immune response. T cell homoeostasis is represented by *s*_*T*_ = *δ*_*T*_ *T* (0), where *T* (0) is the initial number of T cells and *δ*_*T*_ is the half life of T cells. The steady state condition must be satisfied to guarantee the T cell homeostatic value *T* (0) = *s*_*T*_ */δ*_*T*_ in the absence of viral infection.

*K* is the maximum viral load for each of the patients in [21]. The half life of T cells is approximately 4-34 days [39], therefore we take *δ*_*T*_ = 2.9 × 10^− 2^. T cells can proliferate at a rate *r*, and we assumed that the activation of T cell proliferation by *V* follows a log-sigmoidal form with half saturation constant *k*_*T*_. The coefficient *m* relates to the width of the sigmoidal function. While different values of *m* were tested, *m* = 2 rendered a better fit.

Fig.5 show results of parameter fitting for three different scenarios assuming the initiation of the infection (*t*_*i*_) was at -14, -7, and 0 dpso. Panels (a) and (c) of Fig.5 shows that the model (12)-(13) gives a better fitting than previous models (Fig.2-4). Furthermore, AICs values for patient A and B highlight that *t*_*i*_ = − 15 dpso give the best fitting. For presentation purposes, numerical results for patient A and B are the only portrayed in Fig.4. The summary of fitting procedures at *t*_*i*_ = − 15 dpso is presented in Table 3. Independently of the starting infection day, the immune response by T cells peaks between 5 to 10 dpso. Interestingly, the longer the period between infection time to the onset of symptoms, the higher the immune response.

**Fig 5.**
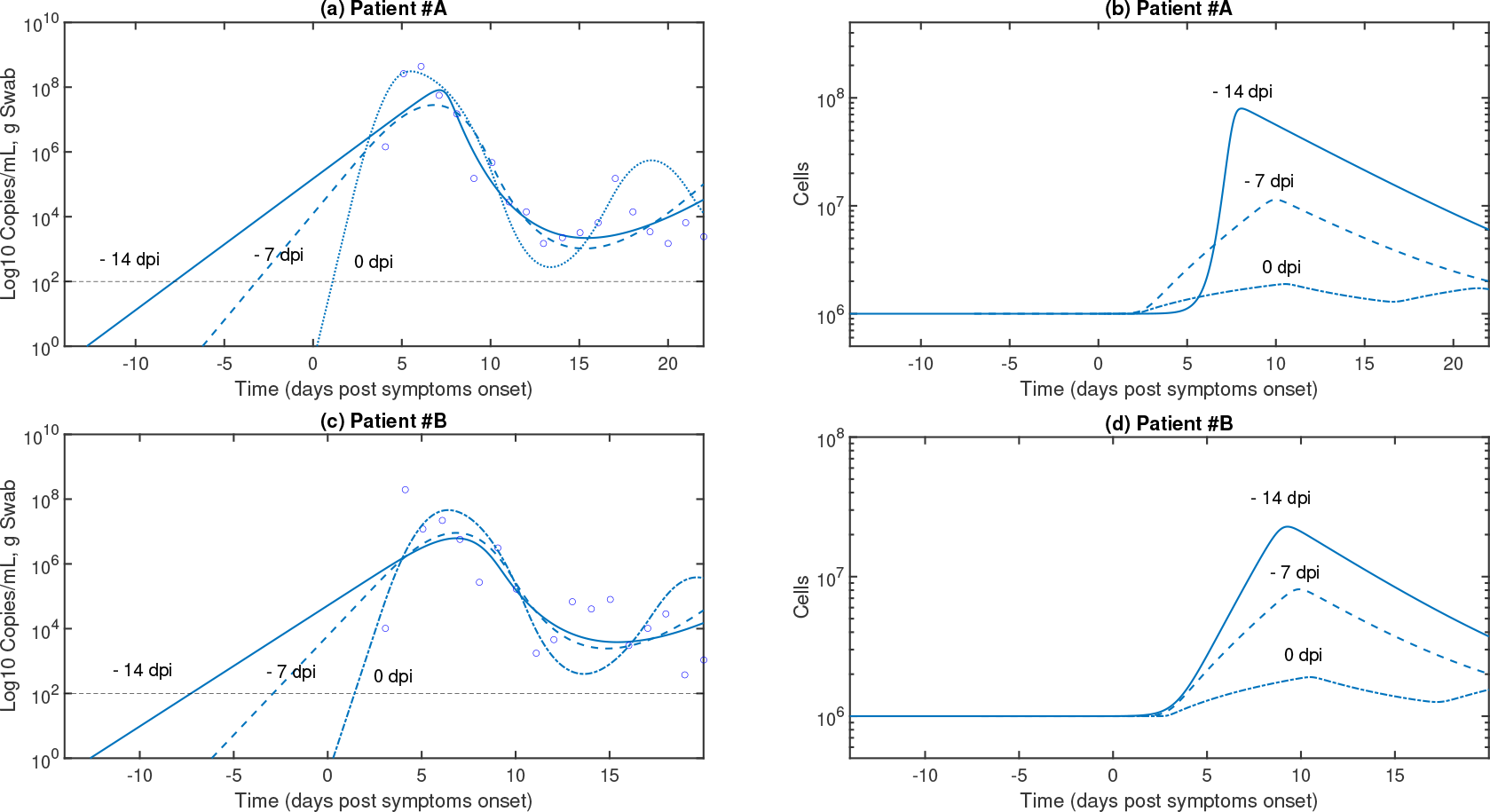
Model with Immune Responses for SARS-CoV-2. Continuous line are simulation based on the target cell model (12)-(13). Blue circles represents the data from [21]. Due to the most complete data sets in [21] were from patient A and B, then these are the only presented in panel (a) and (b), respectively. Infection time was assumed at -14, -7 and 0 days post symptom onset.

**Table 3.**
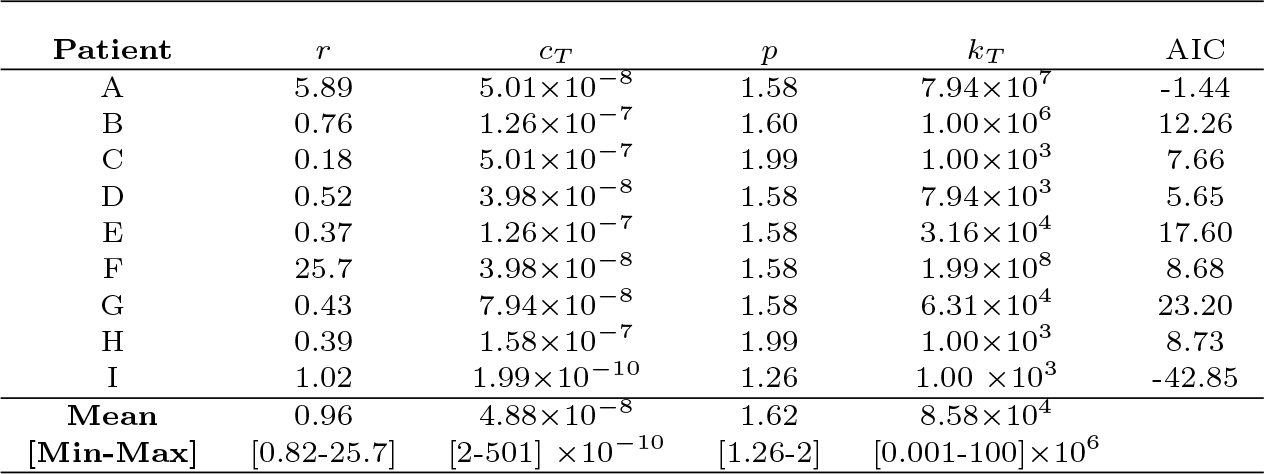
Estimations for model with immune system (12)-(13) using experimental data from [21] assuming *m* = 2 and infection time -14 dpso.

| Patient | $r$ | $c_T$ | $p$ | $k_T$ | AIC |
| --- | --- | --- | --- | --- | --- |
| A | 5.89 | $5.01 \times 10^{-8}$ | 1.58 | $7.94 \times 10^7$ | -1.44 |
| B | 0.76 | $1.26 \times 10^{-7}$ | 1.60 | $1.00 \times 10^6$ | 12.26 |
| C | 0.18 | $5.01 \times 10^{-7}$ | 1.99 | $1.00 \times 10^3$ | 7.66 |
| D | 0.52 | $3.98 \times 10^{-8}$ | 1.58 | $7.94 \times 10^3$ | 5.65 |
| E | 0.37 | $1.26 \times 10^{-7}$ | 1.58 | $3.16 \times 10^4$ | 17.60 |
| F | 25.7 | $3.98 \times 10^{-8}$ | 1.58 | $1.99 \times 10^8$ | 8.68 |
| G | 0.43 | $7.94 \times 10^{-8}$ | 1.58 | $6.31 \times 10^4$ | 23.20 |
| H | 0.39 | $1.58 \times 10^{-7}$ | 1.99 | $1.00 \times 10^3$ | 8.73 |
| I | 1.02 | $1.99 \times 10^{-10}$ | 1.26 | $1.00 \times 10^3$ | -42.85 |
| Mean | 0.96 | $4.88 \times 10^{-8}$ | 1.62 | $8.58 \times 10^4$ | |
| [Min-Max] | [0.82-25.7] | $[2-501] \times 10^{-10}$ | [1.26-2] | $[0.001-100] \times 10^6$ | |

## DISCUSSION

The novel coronavirus SARS-CoV-2 first reported in Wuhan in December 2019 has paralysed our societies, leading to self isolation and quarantine for several days. Indeed, COVID-19 is a major threat to humans, with alarming levels of spread and death tolls, in particular on the eldery. The WHO situation report published on 21 March 2020 reported 267013 confirmed cases and 11201 deaths [1]. COVID-19 is the first pandemic after the H1N1 “swine flu” in 2009 [1]. While many mathematical models have concentrated on the epidemiological level predicting how SARS-CoV-2 would spread, this paper aims to model SARS-CoV-2 dynamics at the within-host level to quantify SARS-CoV-2 infection kinetics in humans.

Data from [26] showed that MERS-CoV levels peak during the second week with a median value of 7.21 (log10 copies/mL)in the severe patient group, and about 5.54 (log10 copies/mL) in the mild group. For SARS, the virus peaked at 5.7 (log10 copies/mL) between 7 to 10 days after onset [40]. For COVID-19, the viral peak was approximately 8.85 (log10 copies/mL) before 5 dpso [21]. Liu *et al*. [41] found that patients with severe disease reported a mean viral load on admission 60 times higher than that of the mean of mild disease cases, implying that higher viral loads relate clinical outcomes. Additionally, higher viral load persisted for 12 days after onset [41].

Using the target cell model, Nguyen *et al*. [13] computed for Ebola infection an average infecting time of 9.49 hours, while Holder *et al*. [29] reported that infecting time for the wild-type (WT) pandemic H1N1 influenza virus was approximately 0.5 hours [29]. Here, based on the results of the target cell model in Table 2, we found that SARS-CoV-2 infecting time between cells (mean of 30 days approximately) would be slower than those reported for Ebola (about 3 times slower) and influenza (60 times slower). The reproductive number for influenza in mice ranges from 1.7 to 5.35 [42], which is consistent with the values reported for COVID-19.

Interestingly, both of our models (the target cell model (3)-(5) and the model with immune response (12)-(13)), when fitted to the patient A data, predicts that the virus can replicate below detection levels for the first 4 dpi. This could be an explanation of why infected patients with SARS-CoV-2 would take from 2-14 dpi to exhibit symptoms.

The model with immune system (Fig.4(b and d)) highlights that the T cell response is slowly mounted against SARS-CoV-2 [4]. Thus, the slow T cell response may promote a limit inflammation levels [42], which might be a reason to the observations during COVID-19 pandemic of the detrimental outcome on French patients that used non-steroidal anti-inflammatory drugs (NADs) such as ibuprofen.

However, so far, there is not any conclusive clinical evidence on the adverse effects by NADs on SARS-CoV-2 infected patients.

The humoral response against SARS-CoV-2 is urgently needed to evaluate the protection to reinfections. A longitudinal study in rhesus monkeys by Bao *et al*. [43] uncovered that infected monkeys presented viral replication at 7 days post-infection (dpi). Significant increase of specific IgG were detected at 14, 21 or 28 dpi. Infected monkeys were re-challenged after specific antibody tested positively and symptoms vanished. Monkeys with re-exposure presented no recurrence of COVID-19, highlighting that protection can be presented to subsequent exposures. Regarding antiviral drugs, Remdesivir treatment has shown a good prophylactic effect during the first 24 hours post MERS-CoV infection in a non-human primate model [44]. Furthermore, benefits has been reported for therapeutic treatment if provided during 12 hours MERS-CoV infection [44]. Our study here mainly addressed T cell responses, therefore, future modelling attempts should be directed to establish a more detailed model of antibody production and cross-reaction [45] as well as *in silico* testing of different antivirals [46].

There are technical limitations in this study that need to be highlighted. The data for SARS-CoV-2 kinetics in [21] is at the onset of symptoms. This is a key aspect that can render biased parameter estimation as the target cell regularly is assumed to initiate at the day of the infection. In fact, we could miss viral dynamics at the onset of symptoms. For example, from throat samples in Rhesus macaques infected with SARS-CoV-2, two peaks were reported on most animals at 1 and 5 dpi [47].

In a more technical aspect using only viral load on the target cell model to estimate parameters may lead to identifiability problems [48–51]. Thus, our parameter values should be taken with caution when parameters quantifications are interpreted to address within-host mechanisms. For the model with immune system, there is not data confrontation with immune response predictions, thus, new measurements on cytokines and T cell responses would uncover new information.

The race to develop the first vaccine to tackle COVID-19 has started with the first clinical trial just 60 days after the genetic sequence of the virus. Modelling work developed in this paper paves the way for future mathematical models of COVID-19 to reveal prophylactic and therapeutic interventions at multi-scale levels [52–57]. Further insights into immunology and pathogenesis of SARS-CoV-2 will help to improve the outcome of this and future pandemics.

## MATERIAL AND METHODS

### Mathematical models

Mathematical models based on Ordinary Differential Equations (ODEs) are solved using the MATLAB library *ode*45, which is considered for solving non-stiff differential equations [58].

### Viral Kinetic Data of Patients Infected with SARS-CoV-2

The clinical data of 9 individuals is from [21]. Due to close contact with index cases and initial diagnostic test before admission, patients were hospitalized in Munich [21]. Viral load kinetics were reported in copies/ml per whole swab for 9 individual cases. All samples were taken about 2 to 4 days post symptoms. Further details can be found in [21].

### Parameter Estimation

Due to the viral load is measured in Log10 scales, parameter fitting is performed minimizing the root mean square (RMS) difference on Log10 scales between the model predictive output 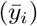, and the experimental measurement (*y*_*i*_):

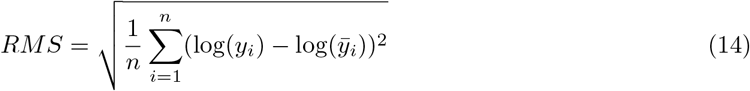

where *n* is the number of measurements. The minimization of RMS is performed using the Differential Evolution (DE) algorithm [22]. Note that several optimization solvers were considered, including both deterministic (*fmincon* Matlab routine) and stochastic (*e.g* Genetic and Annealing algorithm) methods. Simulation results revealed that the DE global optimization algorithm is robust to initial guesses of parameters than other mentioned methods.

### Model Selection by AIC

The Akaike information criterion (AIC) is used here to compare the goodness-of-fit for models that evaluate different hypotheses [59]. A lower AIC value means that a given model describes the data better than other models with higher AIC values. Small differences in AIC scores (*e.g*. <2) are not significant [59]. When a small number of data points, the corrected (AICc) writes as follows:

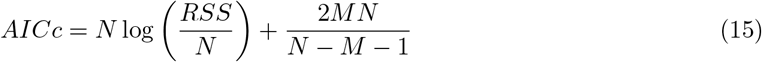

where *N* is the number of data points, *M* is the number of unknown parameters and *RSS* is the residual sum of squares obtained from the fitting routine.

## Data Availability

The origin of the data is clearly stated in the manuscript.

## Author Contributions

EAHV envisaged the project and performed the simulations. All the authors discussed and wrote the paper.

## Acknowledgements

This research was funded by the Universidad Nacional Autonoma de Mexico (UNAM), CONACYT, and the Alfons und Gertrud Kassel-Stiftung. JXVH acknowledges support form grant PAPIIT-UNAM IN115720.

## References

1 CDC. Coronavirus diseases (COVID-2019) situation reports; 2020. Available from: https://www.who.int/emergencies/diseases/novel-coronavirus-2019/situation-reports/.

2 He B, Zhang Y, Xu L, Yang W, Yang F, Feng Y, et al. Identification of diverse alphacoronaviruses and genomic characterization of a novel severe acute respiratory syndrome-like coronavirus from bats in China. Journal of virology. 2014;88(12):7070–82. doi:10.1128/JVI.00631-14.

3 Menachery VD, Yount BL, Debbink K, Agnihothram S, Gralinski LE, Plante JA, et al. A SARS-like cluster of circulating bat coronaviruses shows potential for human emergence. Nature Medicine. 2015;21(12):1508–1513. doi:10.1038/nm.3985.

4 Anderson RM, Heesterbeek H, Klinkenberg D, Hollingsworth TD. How will country-based mitigation measures influence the course of the COVID-19 epidemic? The Lancet. 2020;doi:10.1016/S0140-6736(20)30567-5.

5 Baud D, Qi X, Nielsen-Saines K, Musso D, Pomar L, Favre G. Real estimates of mortality following COVID-19 infection. The Lancet Infectious Diseases. 2020;3099(20):30195. doi:10.1016/s1473-3099(20)30195-x.

6 Li Q, Guan X, Wu P, Wang X, Zhou L, Tong Y, et al. Early Transmission Dynamics in Wuhan, China, of Novel Coronavirus–Infected Pneumonia. New England Journal of Medicine. 2020; p. NEJMoa2001316. doi:10.1056/NEJMoa2001316.

7 Rong L, Ã ASP. Modeling HIV persistence, the latent reservoir, and viral blips. Journal of Theoretical Biology. 2009;260(2):308–331. doi:10.1016/j.jtbi.2009.06.011.

8 Perelson AS, Ribeiro RM. Modeling the within-host dynamics of HIV infection. BMC Biology. 2013;11(1):96. doi:10.1186/1741-7007-11-96.

9 Hernandez-Vargas EA, Middleton RH. Modeling the three stages in HIV infection. Journal of Theoretical Biology. 2013;320:33–40.

10 Pinkevych M, Kent SJ, Tolstrup M, Lewin SR, Cooper DA, Søgaard OS, et al. Modeling of Experimental Data Supports HIV Reactivation from Latency after Treatment Interruption on Average Once Every 5–8 Days. PLoS Pathogens. 2016;12(8):8–11. doi:10.1371/journal.ppat.1005740.

11 Reluga TC, Dahari H, Perelson AS. Analysis if Hepatitis C Virus Infection Models with Hepatocyte Homeostasis. SIAM journal on applied mathematics. 2009;69(4):999–1023.

12 Graw F, Perelson AS. Modeling Viral Spread. Annual Review of Virology. 2015;(July):1–18. doi:10.1146/annurev-virology-110615-042249.

13 Nguyen VK, Binder SC, Boianelli A, Meyer-Hermann M, Hernandez-Vargas EA. Ebola virus infection modeling and identifiability problems. Frontiers in Microbiology. 2015;6:1–11.

14 Nguyen VK, Hernandez-Vargas EA. Windows of opportunity for Ebola virus infection treatment and vaccination. Scientific reports. 2017;7(1):8975.

15 Baccam P, Beauchemin C, Macken Ca, Hayden FG, Perelson AS. Kinetics of influenza A virus infection in humans. Journal of virology. 2006;80(15):7590–9.

16 Handel A, Longini IM, Antia R. Neuraminidase inhibitor resistance in influenza: Assessing the danger of its generation and spread. PLoS Computational Biology. 2007;3(12):2456–2464.

17 Hernandez-Vargas EA, Wilk E, Canini L, Toapanta FR, Binder SC, Uvarovskii A, et al. Effects of aging on influenza virus infection dynamics. Journal of Virology. 2014;88(8):4123–31.

18 Pawelek KA, Dor D, Salmeron C, Handel A. Within-host models of high and low pathogenic influenza virus infections: The role of macrophages. PLoS ONE. 2016;11(2):1–16. doi:10.1371/journal.pone.0150568.

19 Hernandez-Vargas EA. Modeling and Control of Infectious Diseases: with MATLAB and R. 1st ed. London: ELSEVIER Academic Press; 2019.

20 Zou L, Ruan F, Huang M, Liang L, Huang H, Hong Z, et al. SARS-CoV-2 Viral Load in Upper Respiratory Specimens of Infected Patients. New England Journal of Medicine. 2020; p. NEJMc2001737. doi:10.1056/NEJMc2001737.

21 Wölfel R, Corman VM, Guggemos W, Seilmaier M, Zange S, Müller MA, et al. Virological assessment of hospitalized patients with COVID-2019. Nature. 2020; p. 1–10. doi:10.1038/s41586-020-2196-x.

22 Storn R, Price K. Differential Evolution - A simple and efficient adaptive scheme for global optimization over continuous spaces. Journal of Global Optimization. 1997;11(4):341–359. doi:10.1023/A:1008202821328.

23 Perelson AS. Modelling Viral and Immune System Dynamics. Nature Reviews Immunology. 2002;2(1):28–36.

24 Ciupe SM, Heffernan JM. In-host modeling. Infectious Disease Modelling. 2017;2(2):188–202. doi:10.1016/j.idm.2017.04.002.

25 Tyrrell DAJ, Myint SH. Coronaviruses. University of Texas Medical Branch at Galveston; 1996. Available from: http://www.ncbi.nlm.nih.gov/pubmed/21413266.

26 Oh Md, Park WB, Choe PG, Choi SJ, Kim JI, Chae J, et al. Viral Load Kinetics of MERS Coronavirus Infection. New England Journal of Medicine. 2016;375(13):1303–1305. doi:10.1056/NEJMc1511695.

27 Diekmann O, Heesterbeek JAP, Metz JAJ. On the definition and the computation of the basic reproduction ratio R0 in models for infectious diseases in heterogeneous populations. Journal of Mathematical Biology. 1990;28(4):365–382. doi:10.1007/BF00178324.

28 Heffernan JM, Smith RJ, Wahl LM. Perspectives on the basic reproductive ratio. Journal of The Royal Society Interface. 2005;2(4):281–293. doi:10.1098/rsif.2005.0042.

29 Holder BP, Simon P, Liao LE, Abed Y, Bouhy X, Beauchemin CAA, et al. Assessing the in vitro fitness of an oseltamivir-resistant seasonal A/H1N1 influenza strain using a mathematical model. PloS one. 2011;6(3):e14767. doi:10.1371/journal.pone.0014767.

30 Pinilla LT, Holder BP, Abed Y, Boivin G, Beauchemin Caa. The H275Y Neuraminidase Mutation of the Pandemic A/H1N1 Influenza Virus Lengthens the Eclipse Phase and Reduces Viral Output of Infected Cells, Potentially Compromising Fitness in Ferrets. Journal of Virology. 2012;86(19):10651–10660. doi:10.1128/JVI.07244-11.

31 Beauchemin CAA, McSharry JJ, Drusano GL, Nguyen JT, Went GT, Ribeiro RM, et al. Modeling amantadine treatment of influenza A virus in vitro. Journal of Theoretical Biology. 2008;254(2):439–451. doi:10.1016/j.jtbi.2008.05.031.

32 Hancioglu B, Swigon D, Clermont G. A dynamical model of human immune response to influenza A virus infection. Journal of Theoretical Biology. 2007;246(1):70–86.

33 Lee HY, Topham DJ, Park SY, Hollenbaugh J, Treanor J, Mosmann TR, et al. Simulation and prediction of the adaptive immune response to influenza A virus infection. Journal of virology. 2009;83(14):7151–7165. doi:10.1128/JVI.00098-09.

34 Pawelek KA, Huynh GT, Quinlivan M, Cullinane A, Rong L, Perelson AS. Modeling Within-Host Dynamics of Influenza Virus Infection Including Immune Responses. PLoS Comput Biol. 2012;8(6):e1002588.

35 Saenz RA, Quinlivan M, Elton D, MacRae S, Blunden AS, Mumford JA, et al. Dynamics of Influenza Virus Infection and Pathology. Journal of Virology. 2010;84(8):3974–3983.

36 Miao H, Hollenbaugh JA, Zand MS, Holden-Wiltse J, Mosmann TR, Perelson AS, et al. Quantifying the Early Immune Response and Adaptive Immune Response Kinetics in Mice Infected with Influenza A Virus. Journal of Virology. 2010;84(13):6687–6698.

37 Boianelli A, Nguyen VK, Ebensen T, Schulze K, Wilk E, Sharma N, et al. Modeling Influenza Virus Infection: A Roadmap for Influenza Research. Viruses. 2015;7(10):5274–5304.

38 Almocera AES, Nguyen VK, Hernandez-Vargas EA. Multiscale model within-host and between-host for viral infectious diseases. Journal of Mathematical Biology. 2018;77(4):1035–1057. doi:10.1007/s00285-018-1241-y.

39 McDonagh M, Bell EB. The survival and turnover of mature and immature CD8 T cells. Immunology. 1995;84(4):514–20.

40 Peiris JSM, Chu CM, Cheng VCC, Chan KS, Hung IFN, Poon LLM, et al. Clinical progression and viral load in a community outbreak of coronavirus-associated SARS pneumonia: a prospective study. Lancet (London, England). 2003;361(9371):1767–72. doi:10.1016/s0140-6736(03)13412-5.

41 Liu Y, Yan LM, Wan L, Xiang TX, L. A, Liu JM, et al. Viral dynamics in mild and severe cases of COVID-19. The Lancet Infectious Diseases;0(0). doi:10.1016/S1473-3099(20)30232-2.

42 Hernandez-Vargas EA, Colaneri P, Middleton RH. Switching Strategies to Mitigate HIV Mutation. IEEE Transactions on Control Systems Technology. 2014;(1):1–6.

43 Bao L, Deng W, Gao H, Xiao C, Liu J, Xue J, et al. Reinfection could not occur in SARS-CoV-2 infected rhesus macaques. bioRxiv. 2020; p. 2020.03.13.990226. doi:10.1101/2020.03.13.990226.

44 de Wit E, Feldmann F, Cronin J, Jordan R, Okumura A, Thomas T, et al. Prophylactic and therapeutic remdesivir (GS-5734) treatment in the rhesus macaque model of MERS-CoV infection. Proceedings of the National Academy of Sciences. 2020; p. 201922083. doi:10.1073/pnas.1922083117.

45 Hernandez-Mejia G, Hernandez-Vargas EA. Uncovering antibody cross-reaction dynamics in influenza A infections. bioRxiv. 2020; p. 2020.01.06.896274. doi:10.1101/2020.01.06.896274.

46 Hernandez-Mejia G, Alanis AY, Hernandez-Gonzalez M, Findeisen R, Hernandez-vargas EA. Passivity-based Inverse Optimal Impulsive Control for Influenza Treatment in the Host. IEEE Transactions on Control Systems Technology. 2019; p. 1–12.

47 Shan C. Infection with Novel Coronavirus (SARS-CoV-2) Causes Pneumonia in the Rhesus Macaques Sciences. Preprint at Research Square; p. 1–16.

48 Xia X. Estimation of HIV/AIDS parameters. Automatica. 2003;39(11):1983–1988. doi:10.1016/S0005-1098(03)00220-6.

49 Miao H, Xia X, Perelson AS, Wu H. On Identifiability of Nonlinear Ode Models and Applications in Viral Dynamics. SIAM review Society for Industrial and Applied Mathematics. 2011;53(1):3–39.

50 Nguyen VK, Hernandez-Vargas EA. Identifiability Challenges in Mathematical Models of Viral Infectious Diseases. IFAC-PapersOnLine. 2015;48(28):2–7. doi:10.1016/j.ifacol.2015.12.135.

51 Nguyen VK, Klawonn F, Mikolajczyk R, Hernandez-Vargas EA. Analysis of Practical Identifiability of a Viral Infection Model. PLOS ONE. 2016; p. e0167568.

52 Feng Z, Velasco-Hernandez J, Tapia-Santos B, Leite MCA. A model for coupling within-host and between-host dynamics in an infectious disease. Nonlinear Dynamics. 2012;68(3):401–411. doi:10.1007/s11071-011-0291-0.

53 Feng Z, Cen X, Zhao Y, Velasco-Hernandez JX. Coupled within-host and between-host dynamics and evolution of virulence. Mathematical Biosciences. 2015;270:204–212. doi:10.1016/j.mbs.2015.02.012.

54 Handel A, Rohani P. Crossing the scale from within-host infection dynamics to between-host transmission fitness: a discussion of current assumptions and knowledge. Philosophical Transactions of the Royal Society B: Biological Sciences. 2015;370(1675):20140302.

55 Almocera AES, Hernandez-Vargas EA. Coupling multiscale within-host dynamics and between-host transmission with recovery (SIR) dynamics. Mathematical Biosciences. 2019;309(August 2018):34–41. doi:10.1016/j.mbs.2019.01.001.

56 Nguyen VK, Mikolajczyk R, Hernandez-Vargas EA. High-resolution epidemic simulation using within-host infection and contact data. BMC Public Health. 2018;18(1). doi:10.1186/s12889-018-5709-x.

57 Parra-Rojas C, Nguyen VK, Hernandez-Mejia G, Hernandez-Vargas EA. Neuraminidase inhibitors in influenza treatment and prevention–Is it time to call it a day? Viruses. 2018;10(9):454. doi:10.3390/v10090454.

58 Mathworks. Solve nonstiff differential equations — medium order method - MATLAB ode45;. Available from: https://www.mathworks.com/help/matlab/ref/ode45.html.

59 Burnham KP, Anderson DR. Model selection and multimodel inference: a practical information-theoretic approach. Springer Science & Business Media; 2002.

